# Development and validation of the Immune Profile Score (IPS), a novel multi-omic algorithmic assay for stratifying outcomes in a real-world cohort of advanced solid cancer patients treated with immune checkpoint inhibitors

**DOI:** 10.1101/2024.11.05.24316583

**Authors:** Alia Zander, Rossin Erbe, Yan Liu, Ailin Jin, Seung Won Hyun, Sayantoni Mukhopadhyay, Ben Terdich, Mario Rosasco, Nirali Patel, Brett M. Mahon, Kate Sasser, Michelle A. Ting-Lin, Halla Nimeiri, Justin Guinney, Douglas R. Adkins, Matthew Zibelman, Kyle A. Beauchamp, Chithra Sangli, Michelle M. Stein, Timothy Taxter, Timothy A. Chan, Sandip Pravin Patel, Ezra E.W. Cohen

## Abstract

**Background:** Immune checkpoint inhibitors (ICIs) have transformed the oncology treatment landscape. Despite substantial improvements for some patients, the majority do not benefit from ICIs, indicating a need for predictive biomarkers to better inform treatment decisions.

**Methods:** A de-identified pan-cancer cohort from the Tempus multimodal real-world database was used for the development and validation of the Immune Profile Score (IPS) algorithm leveraging Tempus xT (648 gene DNA panel) and xR (RNAseq). The cohort consisted of advanced stage cancer patients treated with any ICI-containing regimen as the first or second line of therapy. The IPS model was developed utilizing a machine learning framework that includes tumor mutational burden (TMB) and 11 RNA-based biomarkers as features.

**Results:** IPS-High patients demonstrated significantly longer overall survival (OS) compared to IPS-Low patients (HR=0.45, 90% CI [0.40-0.52]). IPS was consistently prognostic in PD-L1 (positive/negative), TMB (High/Low), microsatellite status (MSS/MSI-H), and regimen (ICI only/ICI + other) subgroups. Additionally, IPS remained significant in multivariable models controlling for TMB, MSI, and PD-L1, with IPS HRs of 0.49 [0.42-0.56], 0.47 [0.41-0.53], and 0.45 [0.38-0.53] respectively. In an exploratory predictive utility analysis of the subset of patients (n=345) receiving first-line (1L) chemotherapy (CT) and second-line (2L) ICI, there was no significant effect of IPS for time to next treatment on CT in L1 (HR=1.06 [0.85-1.33]). However, there was a significant effect of IPS for OS on ICI in L2 (HR=0.63 [0.46-0.86]). A test of interaction was statistically significant (p<0.01).

**Conclusions:** Our results demonstrate that IPS is a generalizable multi-omic biomarker that can be widely utilized clinically as a prognosticator of ICI based regimens.

**Graphical Abstract:** 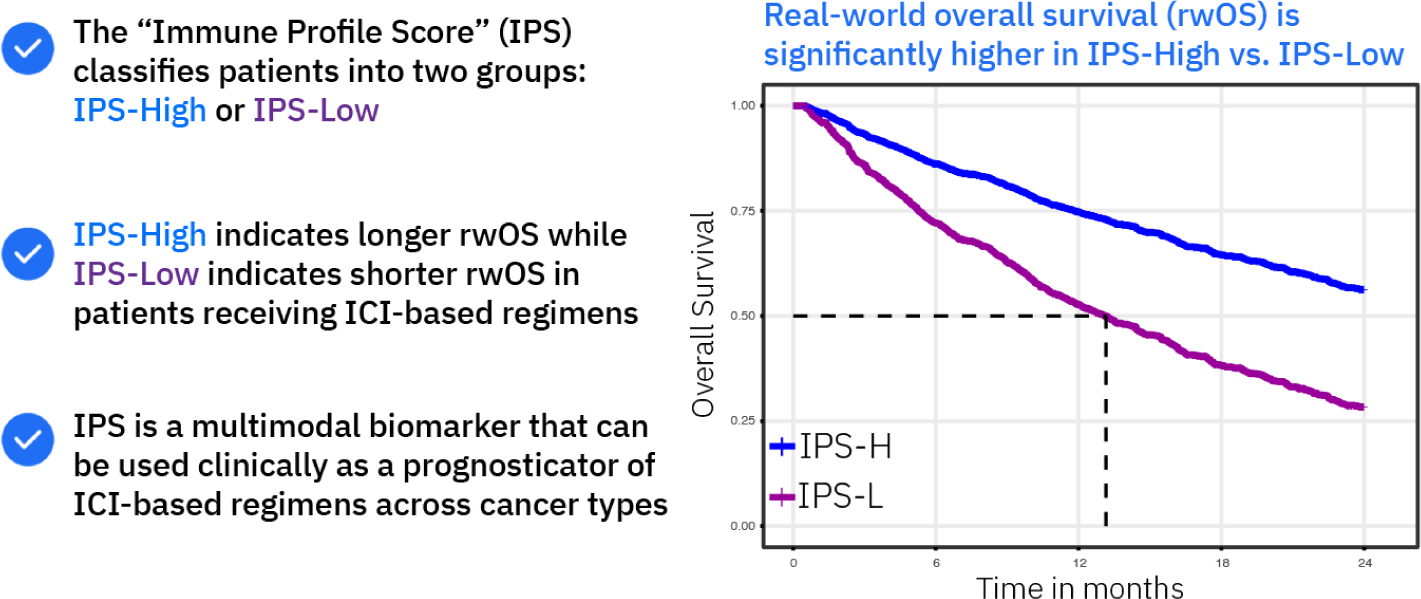

**Key Messages:**

- **What is already known on this topic** – Advancements of multi-omic profiling technology in research settings has demonstrated the potential value of novel immune biomarkers for forecasting response to ICI therapies. However, despite these advances there remains an unmet clinical need for implementation of more sensitive and generalizable biomarkers to better predict patient outcomes to ICI due to limited availability of clinical multi-omic testing and validation cohorts.
- **What this study adds** – Our results demonstrate that IPS is a generalizable multi-omic biomarker that can be widely utilized clinically as a prognosticator of ICI based regimens. Importantly, IPS-High may identify patients within subgroups (TMB-L, MSS, PD-L1 negative) who benefit from ICI beyond what is predicted by existing biomarkers.
- **How this study might affect research, practice or policy** – In the near term IPS results can support patient stratification across pan-solid tumor cohorts to help inform clinicians and researchers which patients are more likely to benefit from ICI based regimens. In the future IPS may support label expansion of ICIs into cancer types without current approvals, and also potentially improve patient selection to minimize over-treatment with ICI in patients unlikely to respond.

## Background

Cancer immunotherapies, particularly immune checkpoint inhibitors (ICIs) targeting PD-[L]1 and CTLA-4, have transformed the oncology treatment landscape. This transformation has been especially notable in cases where conventional systemic therapy options were associated with poor long-term outcomes [1]. Despite substantial improvements, the majority of patients do not benefit from ICIs, emphasizing the need for predictive biomarkers to inform treatment decisions [2].

To date, identifying candidates for immunotherapy relies on myriad PD-L1 immunohistochemistry (IHC) staining criteria across cancer types in addition to pan-cancer biomarkers of microsatellite instability (MSI) status and tumor mutational burden (TMB). Although PD-L1 positivity or high TMB may suggest potential responsiveness to ICIs, there remains a clinical need to improve our ability to determine whether patients will benefit from ICI treatment given the significant number of patients who do not under current guidelines [3].

Translational research efforts have made significant strides in identifying molecular biomarkers beyond PD-L1 IHC, TMB, and MSI, which characterize various aspects of the cancer-immunity cycle that hold promise as predictive immunotherapy biomarkers [4]. Advancements in RNA profiling technologies for both fresh tissue and formalin fixed paraffin embedded tissues have been essential in enabling analysis of routine pathology samples from clinical trials. As evidenced in a comprehensive analysis of publicly available ICI clinical trial data sets, RNA biomarkers are valuable in complementing DNA biomarkers for characterizing ICI response across solid organ cancers [5]. However, while large-panel DNA sequencing often guides treatment decisions in advanced-stage cancers, the clinical utility and routine implementation of RNA sequencing are still emerging, and consequently, RNA sequencing is less frequently available [6]. Additionally, the clinical validation of predictive biomarkers is constrained by limited large-scale multi-omic datasets that include high-quality clinical outcomes data [7]. We developed and validated a multi-omic, pan-solid cancer biomarker using NGS testing, incorporating both DNA and RNA analysis to predict outcomes of ICI therapy.

## Methods

### IPS model development and feature characterization

To develop a biomarker that robustly stratifies outcomes in pan-cancer, solid tumor, metastatic ICI-treated patients, we randomly divided the Tempus ICI cohort into a 1,707 development patient cohort and held out 1,600 patients for clinical validation (**Tables S6** and **S7**). The development cohort was further subdivided into 1,094 patients for feature selection and model training and 613 were reserved for initial model evaluation. Potential features included in the model were drawn from a comprehensive set of RNA and DNA biomarkers that had been previously implicated in tumor-immune biology or associated with IO-related outcomes. We also considered two novel gene signatures developed as a part of this study that characterize expression patterns of tumor-intrinsic immune resistance (see “Model development”).

Candidate model features were selected using a combination of biological plausibility, association with rwOS in publicly available ICI studies, and favorable analytical properties [5–8]. These candidate biomarkers were included in a preliminary multivariate Cox model, stratified by line of therapy. Feature weights were determined using the combined development and evaluation cohorts (n=1,707) and included the following features: TMB [18], expression of *CD74* [19], *CD274* [20], *CD276* [21], *CXCL9* [22], *IDO1* [23], *PDCD1LG2* [20], *SPP1* [22], *TNFRSF5* [24], scIR signature [15], the meta-analysis literature signature, and a gMDSC signature [25] (**Fig.1 a**). The IPS-L and IPS-H thresholds were set as the 55th and 60th percentile of the full training cohort respectively. Patients that fell between the 55th and 60th percentile thresholds were classified as indeterminate and excluded from further analysis.

**Figure 1.**
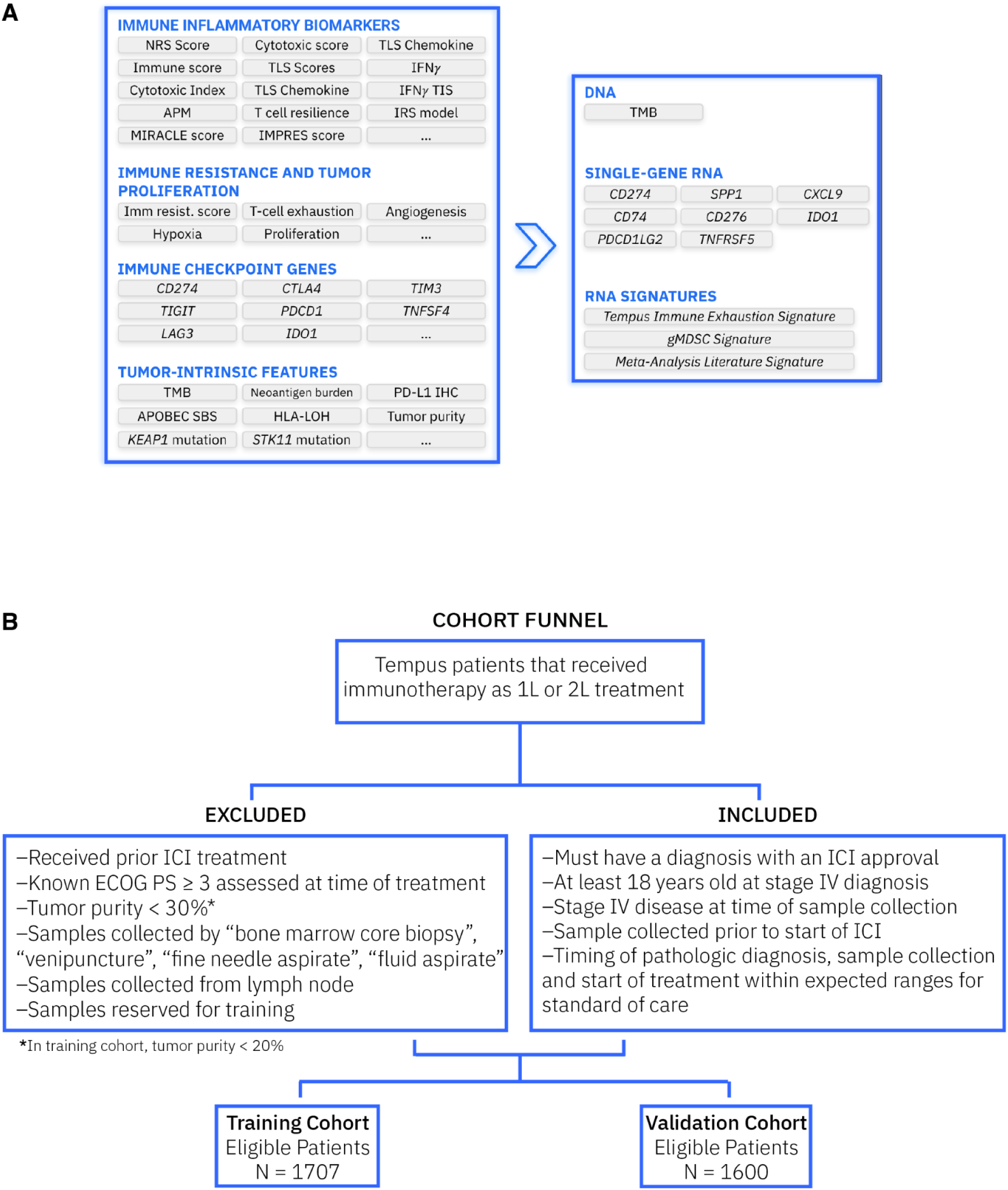
Study Overview. **a)** IPS model features. The Tempus IO Platform was leveraged for developing the IPS model. Various machine learning (ML) techniques were implemented to reduce the feature space when applied to the training and evaluation data set (n=1707). The final IPS model includes 11 RNA-based features and TMB. **b)** Inclusion/exclusion criteria for the training and validation study cohorts.

### Patient Cohorts

Model development and validation cohorts consisted of patients from the de-identified Tempus real-world multimodal database, all of whom underwent clinical next-generation sequencing. **Figure 1b** illustrates the CONSORT diagram for the validation cohort. Patients included in the study were diagnosed with stage IV cancer and received an approved ICI (**Table S1**) in 1L or 2L therapy after January 1, 2018 and before July 1, 2023 (1L) or January 1, 2024 (2L). Cancer types included in the validation and development cohorts are listed in **Tables S2** and **S3**. Patients with an ECOG score ≥ 3 were excluded. To be eligible, samples had to be collected prior to any exposure to ICI therapy, with the time between sample collection and treatment within the standard of care range. Exclusion criteria included low tumor purity (<20% for development, <30% for validation) and samples collected from cytology or lymph node biopsies due to ambiguity of anatomic location of lymph node biopsy, high expression of immune genes in the lymph node, and background noise. Eligible patients were then representatively divided into development (n=1707) and validation (n=1600) cohorts. Overall validation cohort characteristics are listed in **Table 1**.

**Table 1.** Clinical Validation Cohort Characteristics.

| Characteristics | Overall<br>N=1600 | IPS-High<br>N=576 | IPS-Low<br>N=943 | Indeterminate<br>N=81 |
| --- | --- | --- | --- | --- |
| **Age**<br>Mean (SD)<br>Median<br>[IQR]<br>Min / Max | 64.6 (11.8)<br>65.0<br>[58.0, 73.0]<br>20.0 / 89.0 | 64.9 (12.1)<br>65.0<br>[59.0, 74.0]<br>27.0 / 89.0 | 64.5 (11.6)<br>65.0<br>[58.0, 73.0]<br>20.0 / 88.0 | 64.2 (11.7)<br>66.0<br>[56.0, 74.0]<br>34.0 / 83.0 |
| **Sex**<br>Female<br>Male | 645 (40%)<br>955 (60%) | 252 (44%)<br>324 (56%) | 360 (38%)<br>583 (62%) | 33 (41%)<br>48 (59%) |
| **Line of therapy**<br>1<br>2 | 1,326 (83%)<br>274 (17%) | 482 (84%)<br>94 (16%) | 774 (82%)<br>169 (18%) | 70 (86%)<br>11 (14%) |
| **Treatment regimen**<br>**IO Only**<br>IO mono<br>IO doublet<br>**IO + Other**<br>IO+Chemo<br>IO+Chemo+Other<br>IO+Other | 534 (33%)<br>381 (24%)<br>153 (9.6%)<br>1,066 (67%)<br>869 (54%)<br>72 (4.5%)<br>125 (7.8%) | 215 (37%)<br>146 (25%)<br>69 (12%)<br>361 (63%)<br>283 (49%)<br>15 (2.6%)<br>63 (11%) | 292 (31%)<br>219 (23%)<br>73 (7.7%)<br>651 (69%)<br>542 (57%)<br>55 (5.8%)<br>54 (5.7%) | 27 (33%)<br>16 (20%)<br>11 (14%)<br>54 (67%)<br>44 (54%)<br>2 (2.5%)<br>8 (9.9%) |
| **ECOG**<br>0<br>1<br>2<br>Unknown/Missing | 334 (21%)<br>473 (30%)<br>140 (9%)<br>653 (40%) | 137 (24%)<br>168 (29%)<br>45 (8%)<br>226 (39%) | 186 (20%)<br>279 (30%)<br>93 (10%)<br>385 (40%) | 11 (14%)<br>26 (32%)<br>2 (2%)<br>42 (52%) |
| **Stage at primary dx**<br>Stage I<br>Stage II<br>Stage III<br>Stage IV<br>Unknown/Missing | 47 (3%)<br>68 (4%)<br>94 (6%)<br>1,219 (76%)<br>172 (11%) | 22 (4%)<br>25 (4%)<br>33 (6%)<br>430 (75%)<br>66 (11%) | 23 (2%)<br>41 (4%)<br>58 (6%)<br>721 (76%)<br>100 (11%) | 2 (2%)<br>2 (2%)<br>3 (4%)<br>68 (84%)<br>6 (7%) |
| **Cancer type \***<br>Breast<br>Colorectal cancer<br>Gastroesophageal<br>Hepatocellular<br>HNSCC<br>Melanoma<br>NSCLC<br>RCC<br>Urothelial<br>Other | 86 (5%)<br>46 (3%)<br>171 (11%)<br>40 (2%)<br>125 (8%)<br>102 (6%)<br>647 (40%)<br>131 (8%)<br>137 (9%)<br>115 (7%) | 40 (47%)<br>27 (59%)<br>26 (15%)<br>16 (40%)<br>35 (28%)<br>56 (55%)<br>248 (38%)<br>69 (53%)<br>36 (26%)<br>23 (20%) | 41 (48%)<br>18 (39%)<br>134 (78%)<br>22 (55%)<br>86 (69%)<br>42 (41%)<br>367 (57%)<br>49 (37%)<br>95 (69%)<br>89 (77%) | 5 (6%)<br>1 (2%)<br>11(6%)<br>2 (5%)<br>4 (3%)<br>4 (4%)<br>32 (5%)<br>13 (10%)<br>6 (4%)<br>3 (3%) |
| **Brain metastases documented** | 265 (17%) | 107 (19%) | 143 (15%) | 15 (19%) |
| **Liver metastases documented** | 362 (23%) | 94 (16%) | 252 (27%) | 16 (20%) |
| **PD-L1 by IHC**<br>Negative<br>Positive<br>Unknown/Missing | 495 (31%)<br>637 (40%)<br>468 (29%) | 149 (26%)<br>250 (43%)<br>177 (31%) | 321 (34%)<br>353 (37%)<br>269 (29%) | 25 (31%)<br>34 (42%)<br>22 (27%) |
| **TMB**<br>High<br>Low | 430 (27%)<br>1,170 (73%) | 250 (43%)<br>326 (57%) | 160 (17%)<br>783 (83%) | 20 (25%)<br>61 (75%) |
| **MSI**<br>High<br>Stable<br>Undetermined | 80 (5.0%)<br>1517 (95%)<br>3 (0.2%) | 45 (7.8%)<br>531 (92%)<br>0 (0%) | 31 (3.3%)<br>909 (96%)<br>3 (0.3%) | 4 (4.9%)<br>77 (95%)<br>0 (0%) |

\*Overall category sums down column; IPS-H, IPS-L, and Indeterminate sum across rows

### NGS-based DNA and RNA sequencing

The Tempus testing platform includes both a targeted DNA sequencing assay (xT), and an exome capture RNA sequencing assay (xR) [8–10]. The current xT assay targets 648 genes, with a panel size of 1.9MB. Prior versions of xT assay, including a 596-gene version and other DNA sequencing assays, were also utilized in the analysis (**Table S4**). TMB was calculated by dividing the number of nonsynonymous mutations by the size of the panel size (**Supplemental Methods**) [11]. xT also probes for loci frequently unstable in tumors with mismatch repair deficiencies, allowing for the assessment of microsatellite instability (MSI) and classifies tumors into MSI-H, and MSS categories [9]. xR is based on the IDT xGen Exome Research Panel v2 backbone, comprising >415K individually synthesized probes and spanning a 34 Mb target region (19,433 genes) of the human genome. Tempus-specific custom spike-in probes are added to enhance target region detection in key areas. Clinically, the xR assay is used for reporting gene fusions, alternative gene splicing, and gene expression algorithms [9,10,12,13].

### PD-L1 Immunohistochemistry

PD-L1 status for each patient was determined by clinical Tempus testing or curated from pathology reports associated with external PD-L1 IHC testing performed at the referring pathology lab. PD-L1-positive and -negative classification for each cancer subtype was defined per the FDA guidelines or clinical trials (**Table S5**). For cancer types lacking established PD-L1 IHC criteria, a generalized threshold of TPS > 1 was used to define positivity; this was also generalizable across PD-L1 clones used in testing.

### Model Development and Machine Learning (ML) and AI Methodologie**s**

DNA and RNA features adapted to the Tempus IO platform were used as the basis for feature selection for the IPS assay. The features in the IO platform consist of a comprehensive list of DNA and RNA-based IO biomarkers, whose association with tumor immune biology and immunotherapy outcomes has been established in other literature [14]. Additionally, two novel gene signatures were developed by Tempus as part of this study. The first is a signature of tumor-intrinsic immune resistance derived from single-cell RNA-sequencing data, which we term the single-cell immune resistance (scIR) signature [15]. Briefly, this signature uses a variational autoencoder to extract biological signal from a single-cell RNA-sequencing sample taken from a lung adenocarcinoma patient. The scIR signature was strongly weighted in a small population of tumor cells within a highly immune-activated tumor environment and included known pathways of immune inhibitory signaling on tumor associated macrophages. The second signature was created to capture known literature meta-analysis signals using 105 genes [16].

From a cohort of 1707 patients treated with ICI, 1094 patients were used to select features for the model; 613 were held out for model evaluation. This train-evaluation split was performed to create comparable cohorts, stratified by line of therapy and cancer type. To avoid overreliance on this training set, candidate features were further evaluated in publicly-available ICI data sets [5–8] using univariate Cox models. Features that did not reach p < 0.05 in any of these datasets were excluded from consideration. Using the remaining features, we fit a multivariate Cox proportional hazards model, stratified by line of therapy (1L or 2L). The model was trained using 10-fold cross-validation, where balanced L1/L2 regularization was applied to remove redundant features, with cross-validation used to determine the regularization weights. The resulting model was then applied to the remaining 613 held-out patients to verify consistent model performance outside of the initial training data. After this assessment, the model’s final feature coefficients were determined from the full 1707 patient training cohort. The IPS was calculated as a linear combination of the coefficients and scaled to fall between 0-100. The threshold for IPS-Low (IPS-L) was set at all patients below the 55th percentile among the full training cohort, IPS-High (IPS-H) at greater than or equal to the 60th percentile, and the patients between the 55th and 60th percentiles form an indeterminate category.

### Statistical analyses

The analyses conducted in this study were defined prospectively. The primary objective was to demonstrate IPS-H patients had longer overall survival compared to IPS-L patients in a pan-cancer cohort. A stratified Cox proportional hazards model was employed for the primary endpoint of overall survival, with adjustment for treatment regimen type (ICI only vs. ICI+additional), and stratification by line of therapy (1L/ 2L). Risk set adjustment was applied in patients where sequencing (study entry) occurred after the initiation of ICI [17]. The significance of the hazard ratio (HR) was evaluated using a one-sided Wald test at a 5% significance level; consequently, the one-sided upper 90% confidence interval is provided for all survival analyses. The primary endpoint was also descriptively evaluated across several subgroups. These subgroups included PD-L1-positive and -negative patients (based on available IHC data), TMB-H/L (<10 mut/Mb / ≥10mut/Mb), age categories (<65 and ≥65), sex (male/female), regimens (ICI only vs. ICI+additional), and cancer types (restricted to those with at least 15 patients in both the IPS groups). For each of these subgroups, a stratified Cox PH model (incorporating risk set adjustment) similar to the one described in the primary endpoint analysis was fit.

### Statement of Ethics

This study was conducted in accordance with HIPAA regulations where applicable, and IRB exempt determinations (Advarra Pro00076072, Pro00072742).

### Data Availability

Deidentified data used in the research were collected in a real-world health care setting and subject to controlled access for privacy and proprietary reasons. When possible, derived data supporting the findings of this study have been made available within the paper and its Supplementary Figures and Tables.

## Results

### Patient characterization of validation cohort

The validation cohort comprised 1600 patients with stage IV cancer: median (IQR) age of 65.0 (58.0-73.0) years, 40% female (n=645), 1,114 (70%) were treated at community-based hospital or medical practices, and 1,043 (65%) were smokers, 1,016 patients (64%) were White (**Table 1, Table S8**). Most patients in the study were *de novo* stage IV at the time of diagnosis (1,219 [76%]). There were 16 cancer types included in the validation study (**Table S2**). The most common cancer was NSCLC (647 patients [40.4%]), followed by GEJ (171 [11%]), urothelial (137 [9%]), RCC (131 [8%]) and HNSCC (125 [8%]) (**Table S9**). The highest rates of IPS-H were observed in colorectal cancer (27 [59%]), melanoma (56 [55%]), and RCC (69 [53%]) subcohorts (**Table 1**). Consistent with current standards of care, 91% of the colorectal cancer patients were MSI-H (**Table S10**). The lowest rates of IPS-H were observed in GEJ (26 [15%]), urothelial (36 [26%]), and HNSCC (35 [28%]). PD-L1 IHC results were available for 1,132 patients (PD-L1 positive - [637], PD-L1 negative - [495]), the vast majority of cases were stained with PD-L1 22c3 (1,145) (**Table S11**). Notably, a higher proportion of IPS-H patients were PD-L1 positive (250 [43%]) versus PD-L1 negative (149 [26%]). TMB data were available on all patients in the study, and a higher proportion (783 [83%]) of IPS-L patients were TMB-L versus TMB-H (160 [17%]).

Of patients treated with FDA-approved ICIs (**Table S1**), the majority received ICI therapy 1L (1,326 [83%]) versus 2L (274 [17%]). Treatment patterns with ICI were generally consistent with established standards of care. Of the ICI regimen types, ICI+chemotherapy (869 [54%]) was the most common, followed by ICI monotherapy (381 [24%]) and ICI doublet (153 [9.6%]). Notable cancer types and regimens include NSCLC (ICI mono - (92 [14%]), ICI doublet (30 [4.6%]) ICI+chemo - (525 [81%]), melanoma (ICI mono - (56 [55%]), ICI doublet - (40 [39%])), and RCC (ICI doublet - (53 [40%]), ICI+other (66 [50%]) (**Table S12**). Of the patients receiving ICI+other, the “other” consisted mainly of tyrosine kinase inhibitors (78 [4.8%]) of which the majority was used in RCC patients (ICI+TKI - [66]), and ICI with a biologic such as anti-VEGF in hepatocellular carcinoma (Biologic + ICI [26]) and anti-EGFR in GEJ (Biologic + Chemo + ICI - [30]) (**Table S13**). The median follow-up time was 21.2 months (IPS-H) or 18.9 months (IPS-L); follow-up time was calculated from reverse Kaplan Meier (**Table S14**).

### Clinical validation of IPS as a pan-cancer ICI biomarker

A multivariate CoxPH controlling for regimen (ICI monotherapy or ICI combined with other therapies), and stratified by line of therapy (1L or 2L), was used to assess the prognostic association of IPS-H and IPS-L labels with patient outcomes. OS was significantly longer in patients with tumors classified as IPS-H vs IPS-L (HR=0.45 [0.40, 0.52], p-value < 0.01) (**Fig.2 a,b; e**). Differences in survival between IPS-H and IPS-L were consistent across lines of therapy and regimens. The predicted OS from the CoxPH model is shown in **Figure 2** for the setting of ICI-only in 1L (**Fig.2 c**) or 2L (**Fig.2 d**). Notably, the predicted OS curves for ICI combination therapy in 1L and 2L demonstrate a similar relationship to the IPS result and predicted OS. As an exploratory analysis, we assessed the prognostic association of IPS as a continuous variable. The HR (for a 50 unit increase, HR=0.29 [0.24, 0.35]) was similar in magnitude and direction to the categorical representation of IPS.

**Figure 2.**
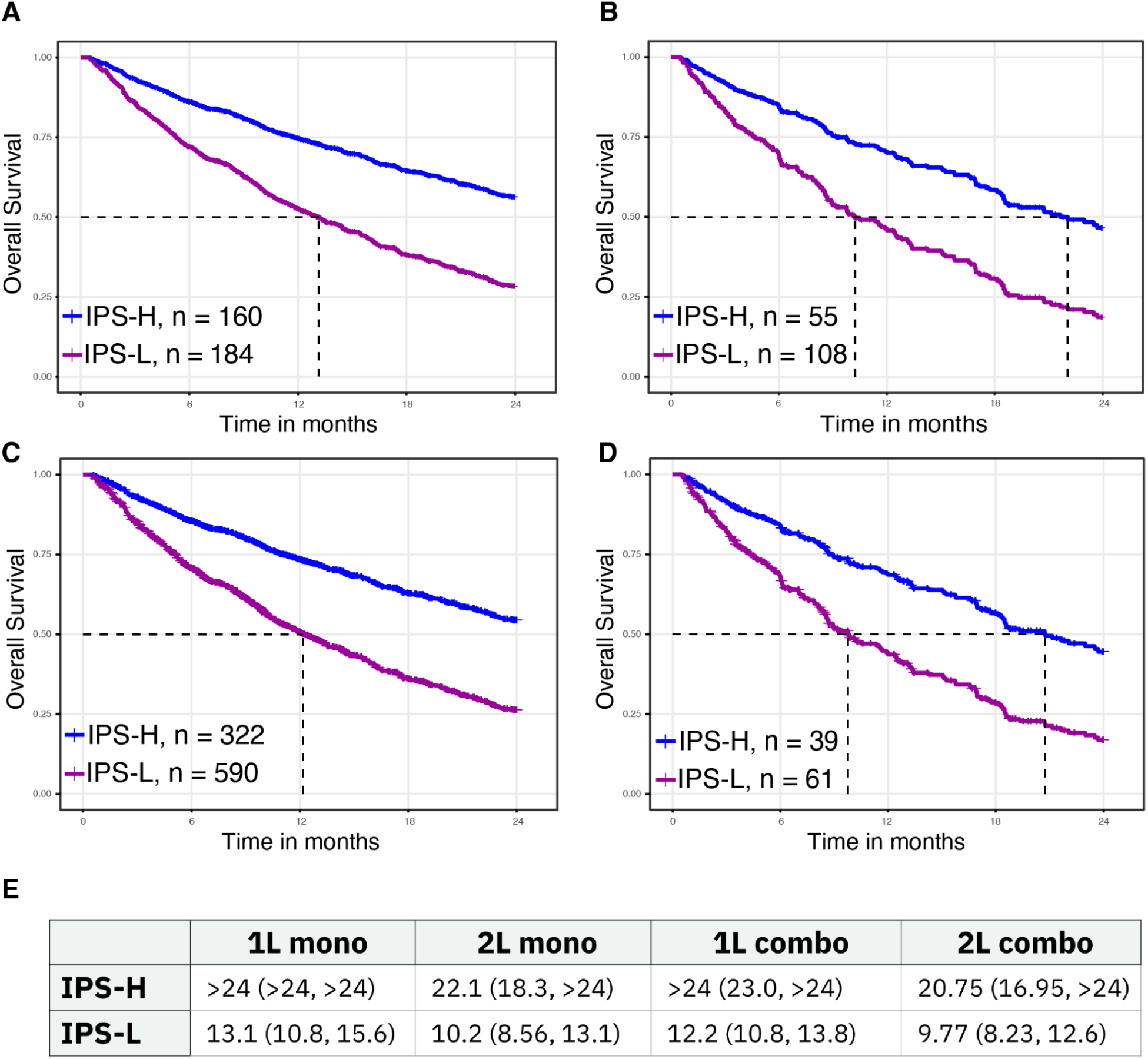
The HR was 0.45 (0.40, 0.52), p < 0.01. Predicted OS from the CoxPH model for **a)** 1L monotherapy and **b)** 2L monotherapy patients. Predicted survival for 1L and 2L combination therapy patients are similar to above. **c)** Predicted OS curves from the CoxPH model in 1L combination therapy (shown in **a**). **d)** Predicted OS curves from the CoxPH model in 2L combination therapy patients (shown in **b**). **e)** The median OS and 95% confidence interval for IPS-H and IPS-L groups for each line of therapy/treatment group combination.

### Performance of IPS in clinical and biomarker subgroups

Prognostic association of IPS in clinical and biomarker subgroups showed that patients whose tumors were classified as IPS-H had significantly longer OS than IPS-L tumors across all subgroups. Notably, significant associations were maintained across molecular biomarker subgroups of TMB-H, TMB-L, PD-L1+, PD-L1-, and MSS as well as clinical subgroups of presence/absence of brain or liver metastasis (**Fig.3**). HR subgroup estimates were similar in direction and magnitude to that of the overall estimate. Among the cancer subgroups evaluated, RCC (0.34 [0.20, 0.59]), HNSCC (0.38 [0.22, 0.67]), NSCLC (0.42 [0.34, 0.52]), and melanoma (0.47 [0.27, 0.82]) had the largest effects while the smallest effects for the IPS were observed in GEJ, HCC, breast and colorectal cancer (CRC). An exploratory subgroup analysis was performed in melanoma (**Fig.S1 a,b**) and RCC (**Fig.S1 c,d**) to evaluate IPS in patients receiving ICI-doublet regimens which are enriched in those disease groups, with melanoma showing (HR=0.56 [0.25-1.23]) and RCC (HR=0.25 [0.10-0.63]).

**Figure 3.**
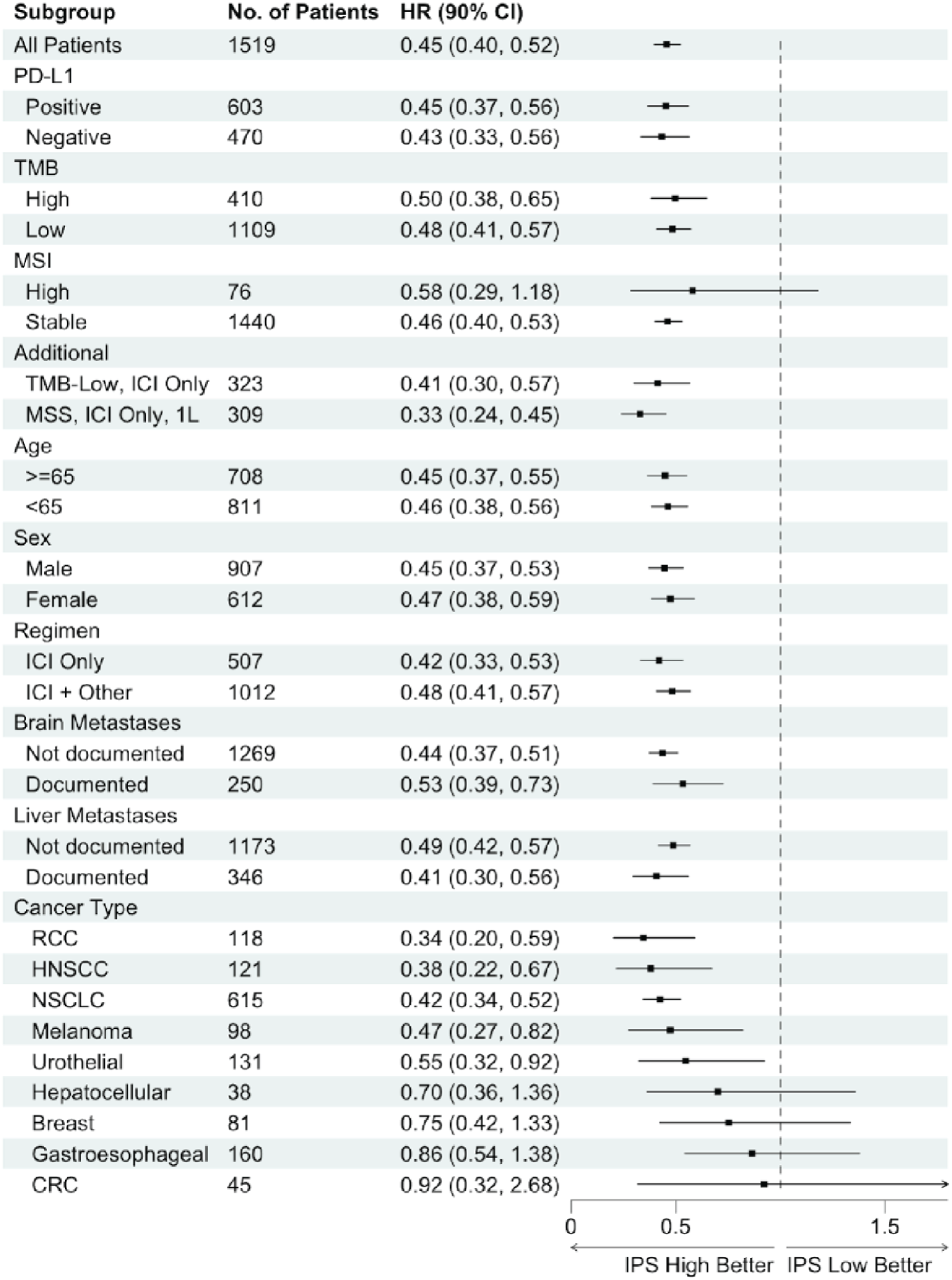
Forest plot showing IPS-H vs. IPS-L hazard ratios and confidence intervals across demographics and clinically relevant subgroups. Subgroups may have <1519 patients due to availability of data.

### IPS complements TMB, PD-L1, and MSI in forecasting responses to ICI

To demonstrate the complementary and added value of IPS in relation to clinically established biomarkers (TMB, PD-L1 IHC, MSI), we compared the full model including both IPS and the biomarker of interest to a reduced model of either TMB, PD-L1 IHC, or MSI without IPS. In this analysis, we observed a significant association of IPS over TMB, PD-L1 IHC, and MSI (p < 0.001).

The predicted OS curves for these biomarker subgroups, categorized by IPS status, are presented for patients treated with ICI monotherapy in 1L (**Fig.4 b-d**). Similar predicted OS curves for treatment conditions and lines of therapy are shown in the **Supplement** (**Fig.S2**). Given the size and clinical significance of the NSCLC cohort, these results are also broken out for NSCLC by PD-L1 status. The predicted OS curve is shown for combination therapy in 1L along with similar predicted OS curves for monotherapy and 2L treated patients (**Fig.4 e,g; Fig.S2 j-l; Fig.S3**). HRs and 90% CI for the most relevant curves shown in the predicted OS plots are listed in **Fig.4 f**.

**Figure 4.**
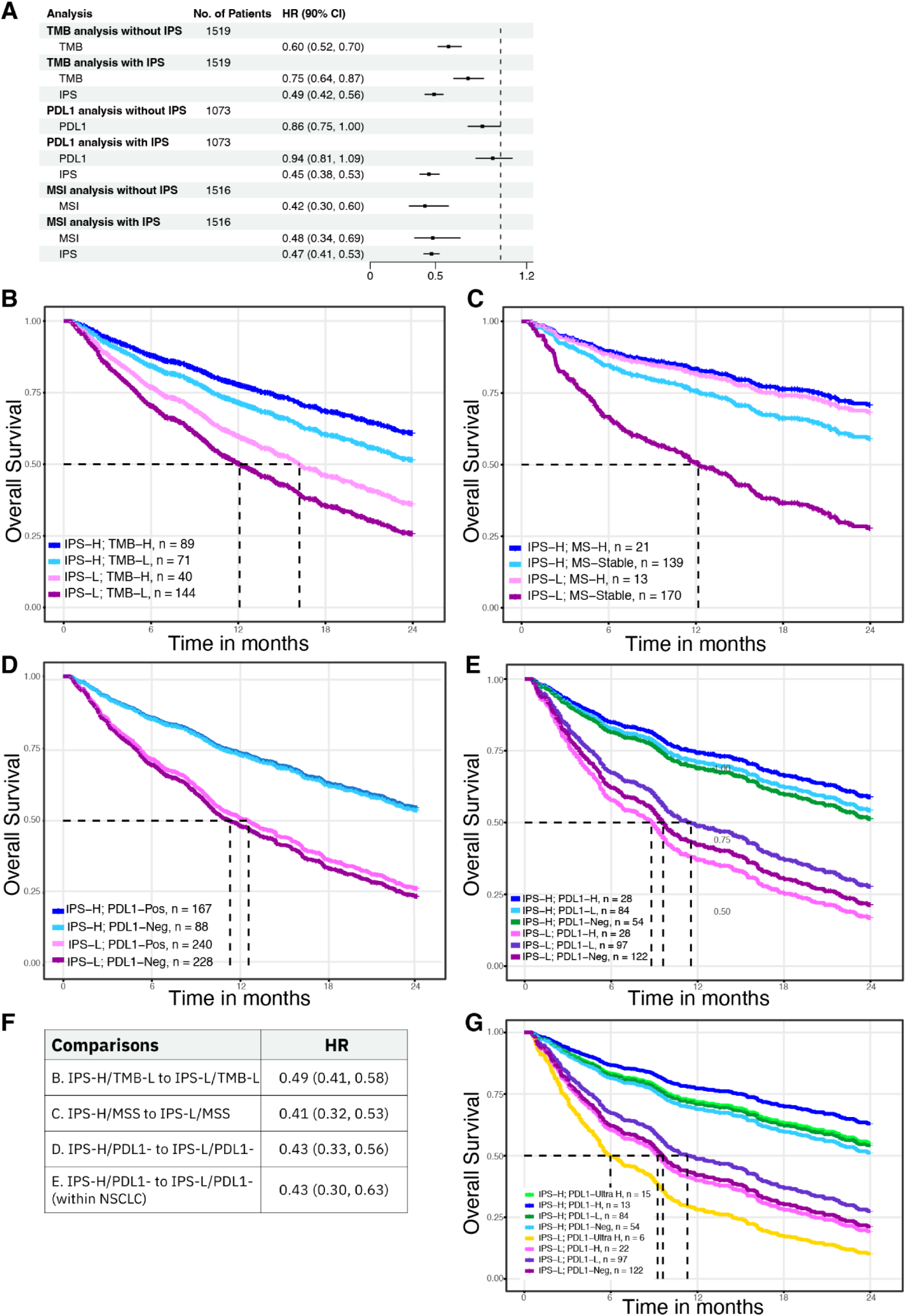
Overall survival is significantly higher in IPS-H vs. IPS-L. **a)** Forest plot showing univariate (UV) HRs for TMB, PD-L1, MSI and multivariate (MV) HRs that include IPS. A likelihood ratio test between the UV and MV models was significant (p<0.01) for all three biomarkers, indicating that IPS has significant prognostic utility beyond TMB, MSI, and PD-L1. Plots **b-e** show predicted OS from a model stratified by line of therapy and fit on IPS, treatment group, and the MV model with the listed biomarker: **b)** TMB pan-cancer, **c)** MSI pan-cancer, **d)** PD-L1 pan-cancer and **e)** PD-L1 in NSCLC patients. The predicted OS curves represent patients treated with monotherapy in 1L for TMB and MSI **(b-c)**, and combination therapy in 1L for PD-L1 and NSCLC **(d,e)**. **f)** HR and 90% CI for the most relevant curves shown in the predicted OS plots in **(b-e)**. **g)** Predicted OS curves from CoxPH Model for NSCLC ICI+combination 1L cohort stratified by IPS result and PDL1 IHC staining level. PD-L1 ultra high: TPS >90, PD-L1 high: TPS=89-50, PD-L1 low: TPS=49-1, PD-L1 negative: TPS=0.

### Exploratory evaluation of predictive utility for IPS

In an exploratory analysis to test the potential predictive utility of IPS, we examined a combined cohort of training and validation patients that had been exposed to non-ICI and ICI therapies in 1L and 2L, respectively. While IPS was not associated with TTNT on CT in 1L (HR=1.06 [0.85, 1.33]; **Fig.5 a**), it was significantly associated with OS in patients receiving 2L ICI (HR=0.63 [0.46, 0.86]; **Fig.5 b**).

**Figure 5.**
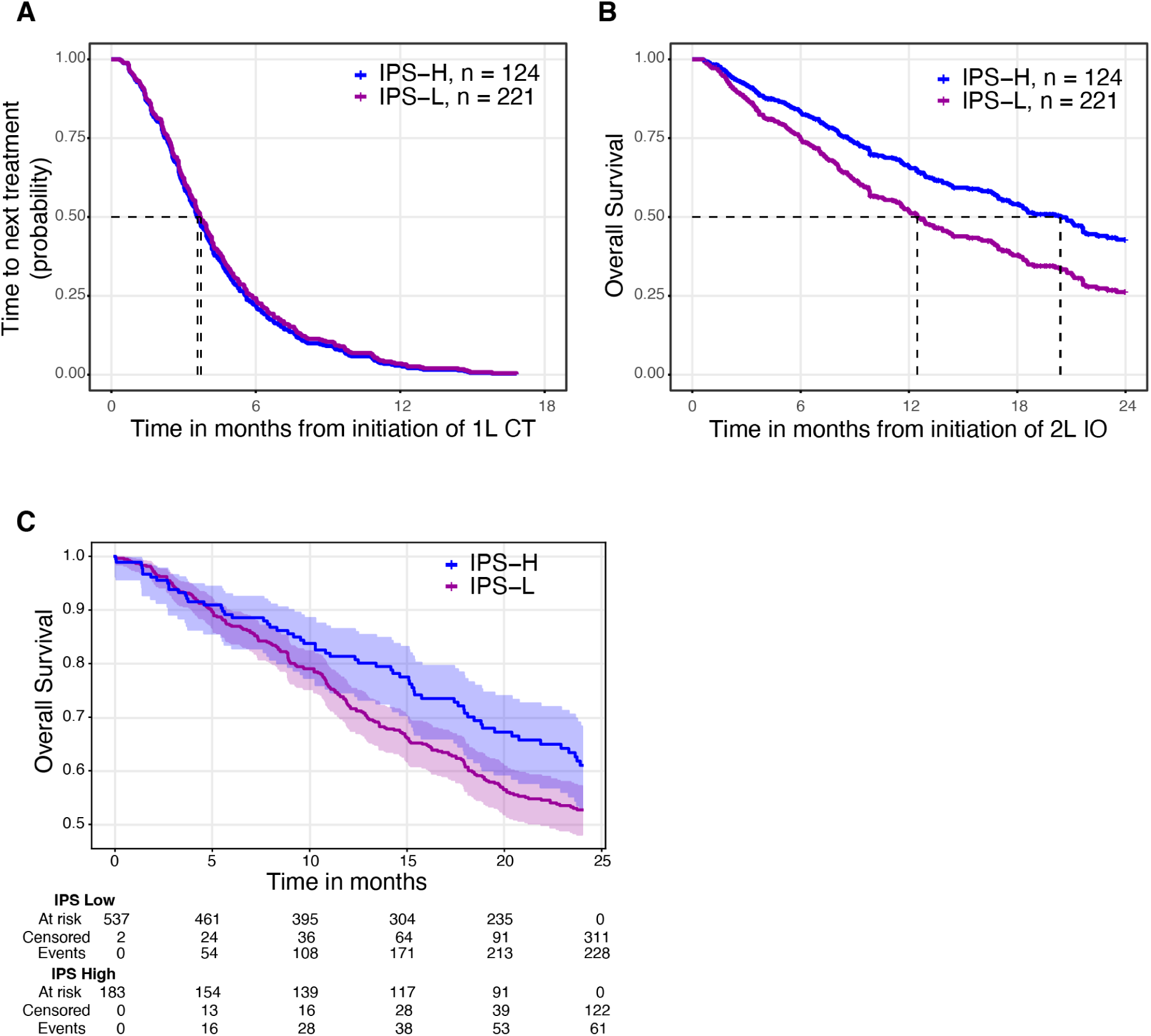
**a)** Predicted TTNT for 1L CT with no significant effect for IPS (HR = 1.06 (0.85, 1.33)). **b)** Predicted OS for 2L ICI shows that IPS does have a significant effect (HR = 0.63 (0.46, 0.86)). Interaction test p < 0.01, indicating that the HR in 2L ICI is significantly different from HR in 1L CT. **c)** KM OS curves for stage IV TCGA patients stratified by IPS result generated from TCGA DNA and RNA data processed through Tempus bioinformatic pipelines.

To further evaluate the prognostic utility of IPS in non-ICI treated patients to understand its predictive utility, we analyzed stage IV patients from The Cancer Genome Atlas (TCGA, N=722, patient selection criteria is described in Supplemental Methods). The TCGA enrollment period was prior to the approval and usage of ICI therapies thus ensuring a representative non-ICI comparator cohort that also had DNA and RNA sequencing available to generate IPS. There was a significant association of IPS with OS in this cohort (HR=0.75 [0.56-0.99]), however the hazard ratio was attenuated relative to the IPS validation cohort. (**Fig.5 c**).

### Tumor distribution and IPS prevalence in an expanded pan-cancer cohort

In order to characterize IPS prevalence more generally, including in cancer types without approved ICI indications, we examined the distribution of IPS-H and IPS-L in an expanded pan-cancer cohort of patients sequenced at Tempus. In the entire cohort encompassing 25 different cancer types, prevalence of IPS-H was 28.64%. Lung adenocarcinoma, RCC, and melanoma had IPS-H prevalence greater than 50% (**Fig.6 a**). On the opposite side of the spectrum, GI neuroendocrine cancer, cholangiocarcinoma, CRC, gynecologic sarcomas, and PDAC all had IPS-H prevalence of less than 20%. Of note, lung squamous cell carcinoma had a prevalence of 25.59% and NSCLC-NOS had a prevalence of 45.75% indicating a likely high proportion of lung adenocarcinomas in the NOS group of patients. To further characterize how IPS may identify ICI responders outside of current cancer type or pan-cancer biomarker ICI approvals, we calculated the proportion of patients who were both IPS-H and TMB-L (14.1%) after excluding cancer types with an ICI approval or tumors that were MSI-H (**Table S15**). We also generated a more granular cancer subtype type visualization of IPS status in relation to TMB status (**Fig.6 b**).

**Figure 6.**
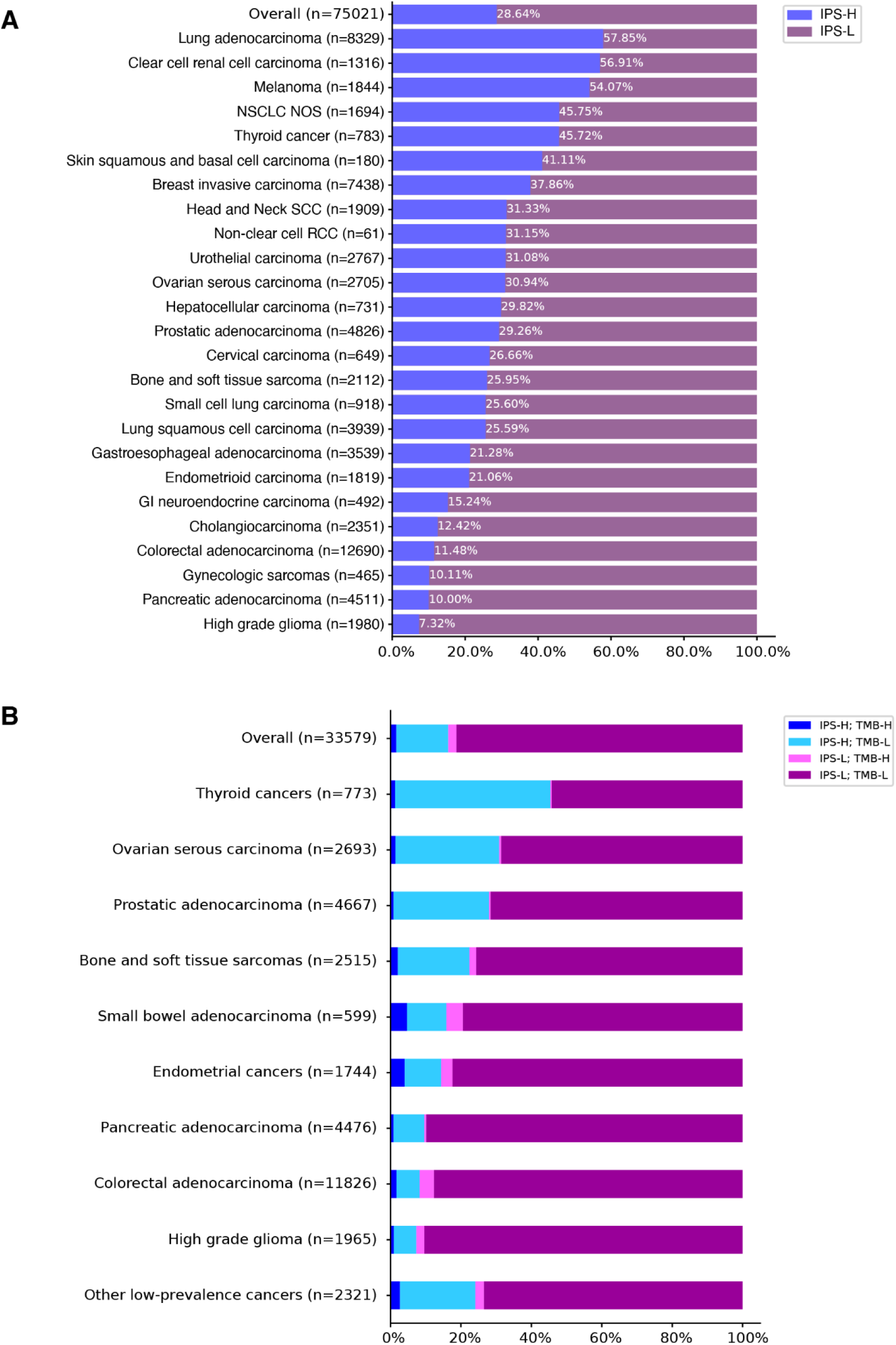
**a)** Prevalence plot showing the percentage of IPS-H patients in a large, representative cohort of Tempus clinical patients. **b)** Prevalence plot showing the percentage of IPS-H and IPS-L patients compared to percentage of TMB-H/L patients in a large, representative cohort of patients from the Tempus multimodal database.

## Discussion

Leveraging the Tempus xT / xR assays and the IO platform along with real-world data from ICI-treated patients, we developed and validated the multi-omic IPS algorithm in a prospectively-designed retrospective study. Using a prespecified statistical analysis plan, IPS was validated as a generalizable pan-cancer prognostic biomarker demonstrating that IPS-H patients have significantly longer OS than IPS-L patients. Additionally, the validation demonstrated that IPS-H patients had significantly longer OS compared to IPS-L patients across relevant ICI biomarker subgroups, including PD-L1 status, TMB levels, and microsatellite stability. Notably, IPS retained its prognostic significance in multivariable models, even when controlling for TMB, MSI status, and PD-L1 expression. Overall, these analyses demonstrate the clinical value of IPS in assessing the potential benefits of ICI regimens beyond current standard of care biomarkers. Finally, a post-hoc exploratory analysis into the predictive capabilities of IPS demonstrated that IPS did not predict TTNT following chemotherapy. However, IPS was a significant predictor of OS when patients were subsequently treated with ICI.

Our study builds upon a growing body of evidence supporting that multi-omic biomarkers developed using machine learning/artificial intelligence methodologies, high-throughput commercial NGS assays, and real-world clinical data can provide insights into tumor/immune biology and clinical outcomes. A vast number of clinical trials (including ALCHEMIST, OptimICE-PCR, EQUATE, PET-Stop trials) utilizing immunotherapies with a diverse range of mechanisms/targets highlights opportunities and unmet clinical needs for patient selection using multi-omic biomarkers [26].

Current paradigms for stage IV solid organ cancers demonstrate opportunities for biomarkers to help inform clinical management for approved ICI regimens in indications with equipoise between regimens or indications that lack biomarkers for patient selection. This is perhaps most apparent in NSCLC where patients of all PD-L1 levels are approved for ICI+chemo while in tumors with PD-L1 IHC high (TPS > 50) patients can receive ICI+chemo or ICI monotherapy [27]. Aguilar *et al.* showed in an RWD retrospective analysis that patients with TPS scores greater than 90 have significantly better outcomes than patients with TPS between 50 and 89, which may be informative for ICI monotherapy patient selection [28]. In our exploratory analysis of NSCLC patients, patients with IPS-H tumors in all PD-L1 IHC subgroups had longer OS than patients with IPS-L tumors. This finding may represent the importance of *CD274* (*PD-L1*) gene expression as a continuous feature in the IPS model. The analysis is notably limited by small sample sizes but generally highlights the potential of IPS to capture tumor immune biomarker stratification and enhance precision. Currently the INSIGNA study which is a large randomized control trial in NSCLC has aims focused on elucidating the optimal clinical management for these patients [29].

IPS may inform new treatment indication strategies. **Figure 6** highlights the cancer-specific IPS-H/L prevalence in an expanded cohort that includes diseases that currently lack ICI indications. Of note, MSS colorectal cancer and pancreatic cancer had among the lowest IPS-H rates consistent with the historically limited response rates seen in ICI monotherapy trials for those cancer types [30–32]. IPS therefore could have value in identifying the rare potential ICI responders in these or similarly challenging diseases. Prospective clinical trial designs utilizing IPS stratification or selection may be considered in the future [33].

Perhaps even more impactful than development of monotherapy ICI studies is the potential application of multi-omic biomarkers such as IPS to inform patient selection for novel ICI combinatorial strategies and the next generation of immunotherapy modalities such as T-cell/NK-cell engagers and RNA cancer vaccines. These novel applications may require modified versions of IPS along with additional biomarkers that characterize the cancer-immunity cycle relevant to a specific combinatorial strategy [4]. Lastly, as ICI based regimens move into neoadjuvant and adjuvant settings, there are significant opportunities for patient selection strategies to reduce ICI exposure in patients unlikely to respond and therefore reducing the number of overall adverse events.

Limitations of this study reflect the real-world, retrospective nature of the validation cohort. While the inclusion and exclusion criteria attempted to control for confounding variables, additional biases may be unaccounted for. Tempus clinical testing and subsequent clinical-molecular data set was generated predominantly in the post ICI-era. Therefore, our predictive analysis did not allow for case-control matching with patients who received non-ICI regimens prior to approvals. We attempted to address this with an analysis of stage IV patients who did not receive ICI, collected from TCGA. Among these patients, we observed a significant difference in OS between patients classified as IPS-H versus IPS-L. This suggests that the IPS model has generalized prognostic utility, as would be biologically expected given the known prognostic association of immune infiltration in tumors [34]. However, given the attenuated hazard ratio we observed in non-ICI-treated patients in TCGA versus the ICI-treated patients in the study cohort, IPS appears to have predictive utility. The disproportionate cancer subgroups are representative of cancer prevalence and NGS testing frequency. The variability of cohort size across cancer types limits our ability to comprehensively evaluate the heterogeneity of IPS performance across cancer types. Additionally, the IPS model did not include clinical and lab features that have been demonstrated to add prognostic utility in combination with molecular markers such as TMB, which could be considered for future model iterations [35].

In summary, we demonstrated in a large RWD clinical validation study that IPS is a generalizable multi-omic biomarker that can be widely utilized clinically as a prognosticator of ICI based regimens. Importantly, IPS-H may identify patients within subgroups (TMB-L, MSS, PD-L1 negative) who benefit from ICI beyond what is predicted by existing biomarkers. Future prospective predictive utility studies are planned for evaluating the clinical applications of IPS.

## List of abbreviations used

IPS: (immune profile score)
ICIs: (immune checkpoint inhibitors)
TMB: (tumor mutational burden)
MSS: (microsatellite stable)
MSI: (microsatellite instability)
OS: (overall survival)
1L/2L: (first/second line)
IHC: (immunohistochemistry)
scIR: (single-cell immune resistance)
TTNT: (time to next treatment)
TCGA: (The Cancer Genome Atlas)

## Acknowledgments

We thank Dana DeSantis from the Tempus Scientific Communications team for writing support. Parts of this work were presented at SITC 2024. A version of this manuscript has been uploaded as a preprint on medRxiv (doi.org/10.1101/2024.11.05.24316583).

