## Supplemental methods and figures for "Development and validation of the Immune Profile Score (IPS), a novel multi-omic algorithmic assay for stratifying outcomes in a real-world cohort of advanced solid cancer patients treated with immune checkpoint inhibitors"

**Supplemental Material**

#### Supplemental Methods

##### *Clinical Data Abstraction*

Clinical data were extracted from the Tempus real-world oncology database. This encompassed longitudinal structured and unstructured data from geographically diverse oncology practices, including integrated delivery networks, academic institutions, and community practices. Structured data from electronic health record systems were integrated with unstructured data collected from patient records via technology-enabled chart abstraction and corresponding molecular data, if applicable. Patients with no recorded date of death across all mortality sources were censored at the date of last recorded interaction with the medical system (i.e., date of last follow-up).

##### *Additional Statistical Analyses*

The prognostic utility of IPS over PD-L1 was evaluated by a likelihood ratio test that compared the full Cox model including both PD-L1 and IPS to a reduced Cox model that included PD-L1 alone (**Methods - Statistical analysis**). The prognostic utility of the IPS in relation to TMB and MSI-H was assessed using a similar approach.

An exploratory analysis of the predictive utility of IPS was performed by combining the training and validation cohorts of patients who received chemotherapy (CT) as first line treatment and ICI as second line treatment. Patients served as their own control in this analysis, and outcomes were evaluated for two lines of therapy: time to next treatment (TTNT) on CT and OS on ICI. If IPS was purely prognostic, time to next treatment (as a surrogate for progression) would be anticipated to be longer in IPS-H patients than in IPS-L patients. The HR for TTNT of IPS-H to IPS-L would then be of a similar magnitude as the HR for OS on second line treatment with ICI.

A conditional model for recurrent events was fit to the selected subset of patients. Specifically, a Cox proportional hazards model, stratified by line of therapy, was used to model the two ordered time periods: period 1 in which the patient received CT and period 2 in which the patient received ICI. A Wald test p-value of less than 0.05 for the interaction between IPS and line of treatment would indicate a significant difference in the hazard ratios between the two time periods.

***TMB***

TMB was calculated by dividing the number of nonsynonymous variations by the size of the panel (2.4 Mb for the panel size of xT.v2 and 1.9Mb for the panel coding region of xT.v4). All non-silent somatic coding variations such as missense, indel, and stop-loss variants with coverage greater than × 100 and an allelic fraction greater than 5% are included in the count of nonsynonymous variations. TMB calculated using the assay is highly correlated with TMB calculated from whole exome TCGA data (R=0.986, P < 2.2 × 10-16). The xT.v2 TMB score is adjusted for differences in denominators between the versions to be directly comparable to xT.v4. All analyses are completed incorporating both assays, with tumors considered TMB-H if they have an adjusted TMB score of 10 mut/Mb or more.

***TCGA Analyses***

FASTQ files from RNA sequencing data for the TCGA cohort were downloaded from the Genomic Data Commons [36] and processed through the Tempus RNA pipeline as described below. The clinical data for the cohort was obtained from cBioportal [37]. Patients included were required to have been Stage 4 at sample collection and have RNA-sequencing, TMB, and OS data available. The period from OS anchor date to the 24 month maximum follow up date was required to be before first FDA approval of ICI. These criteria yielded a cohort of 752 patients. The RNA-sequencing data was reprocessed from raw abundance files using the Tempus

RNA-processing pipeline (as described in **Methods**). Linear batch correction was further applied so that the normalized counts were comparable to the data used in the IPS validation cohort. The IPS model was run on the resulting data set without adjustment. Of the 752 patients, 722 were assigned an IPS-High (IPS-H) or IPS-Low (IPS-L) category, with 30 receiving a score in the indeterminate range.

##### *Analytical Validation*

The Tempus IPS assay was analytically validated to ensure consistent performance across a variety of experimental conditions associated with the underlying IPS assay inputs (xT - TMB, xR - RNA features) to test the precision and analytical accuracy of computing the IPS and the IPS result (IPS high or IPS low) under CAP/CLIA standards. Precision was tested through repeatability and reproducibility studies using tumor samples from five cancer types: NSCLC, HNSCC, melanoma, urothelial carcinoma, and RCC. These samples were run in triplicate, incorporating both DNA (xT) and RNA (xR) replicates to generate an IPS. Repeatability was evaluated within a single assay run, while reproducibility was tested across multiple runs involving different instruments, reagent lots, and operators. 30 tumor samples were used in replicates, with DNA and RNA extracted from the same patients but placed on separate plates for independent processing in each run. The study utilized 12 different flowcell reagent lots, 20 unique flowcells, and 11 distinct sequencers, 4 different operators, ensuring a comprehensive evaluation of reproducibility across different conditions. The repeatability overall percent agreement (OPA) was calculated to be 97% (**Fig.S4 a**), while the reproducibility OPA was determined to be 95% **Fig.S4 b**). Both demonstrated tight clustering of replicate IPS around the expected diagonal, with over 95% of replicate pairs producing highly consistent IPS, affirming the assay's robust repeatability and reproducibility across varied experimental conditions.

Analytic sensitivity was assessed by testing RNA inputs of 25 ng, 50 ng, 100 ng, and 300ng to ensure robust performance at varying RNA levels (**Fig.S4 a,b**). In addition, retrospective real-world data from clinically sequenced samples using the xT and xR assays (the IPS RWD cohort) were used for validation of reportable range and analytic sensitivity, leveraging the combined DNA and RNA features to ensure the assay's reliable performance across diverse tumor sites, procedures, and tumor purity thresholds. New prospective data generated in the wet lab were used in precision experiments and in laboratory concordance studies, ensuring that the IPS assay produces consistent and reproducible results across all solid tumors.

Lastly, we tested the effect of macrodissection and changes in tumor purity on IPS results given the practice of pathologist discretionary macrodissection in the xT and xR sample workflow. Unstained slides from 29 samples that were previously macrodissected as part of the clinical workflow underwent whole slide scraping, resulting in non-macrodissected samples with lower tumor purity than the original microdissected samples. These samples had tumor purities ranging from 40% to 80%, representing borderline macrodissectable cases. A comparison of pre- and post-macrodissected samples (**Fig.S4 e**) revealed a Pearson correlation coefficient of 0.979 and an overall percent agreement in risk classification of 85.7%, indicating a very strong positive correlation between IPS before and after macrodissection. This suggests that the macrodissection process does not significantly impact the assay's results. The robustness of the IPS assay was further confirmed by consistent risk classification across various cancer types, ensuring its reliability for clinical decision-making even in samples with borderline tumor purity.

#### Supplemental Figures

#### Figure S1

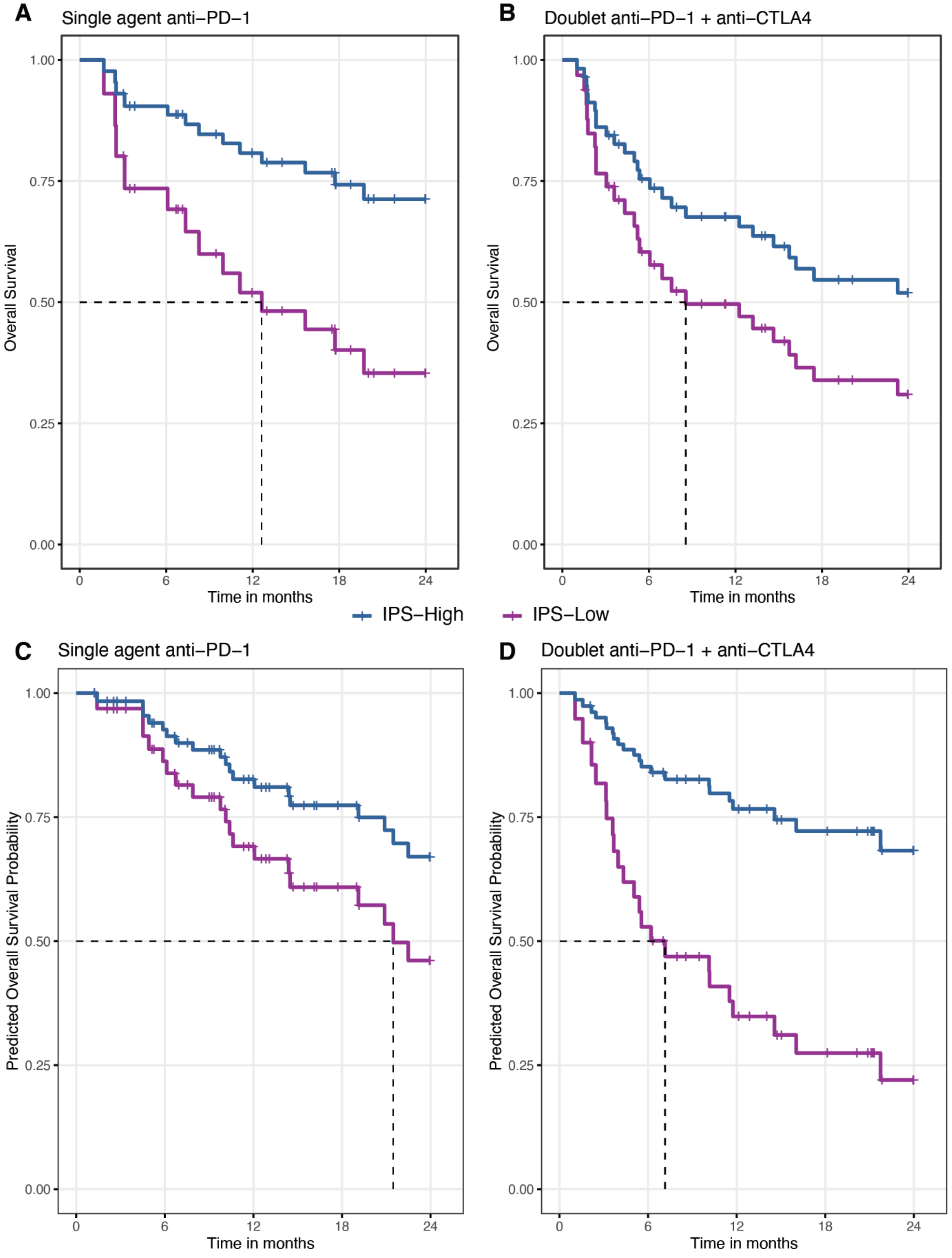
A
Single agent anti−PD−1
B
Doublet anti−PD−1 + anti−CTLA4
C
Single agent anti−PD−1
D
Doublet anti−PD−1 + anti−CTLA4
Overall Survival
Predicted Overall Survival Probability
1.00
0.75
0.50
0.25
0.00
0
6
12
18
24
Time in months
IPS−High
IPS−Low

**Figure S1.** Predicted OS curves for a univariate CoxPH model fit on IPS in melanoma patients treated with **a)** single anti-PD-1 agents (IPS-H, n=26; IPS-L, n=17) and **b)** doublet anti-PD-1 and anti-CTLA1 agents (IPS-H, n=30; IPS-L, n=25). Predicted OS curves for a univariate CoxPH model fit on IPS in RCC patients treated with **c)** single anti-PD-1 agents (IPS-H, n=41; IPS-L, n=26) and **d)** doublet anti-PD-1 and anti-CTLA1 agents (IPS-H, n=28; IPS-L, n=23).

### Figure S2

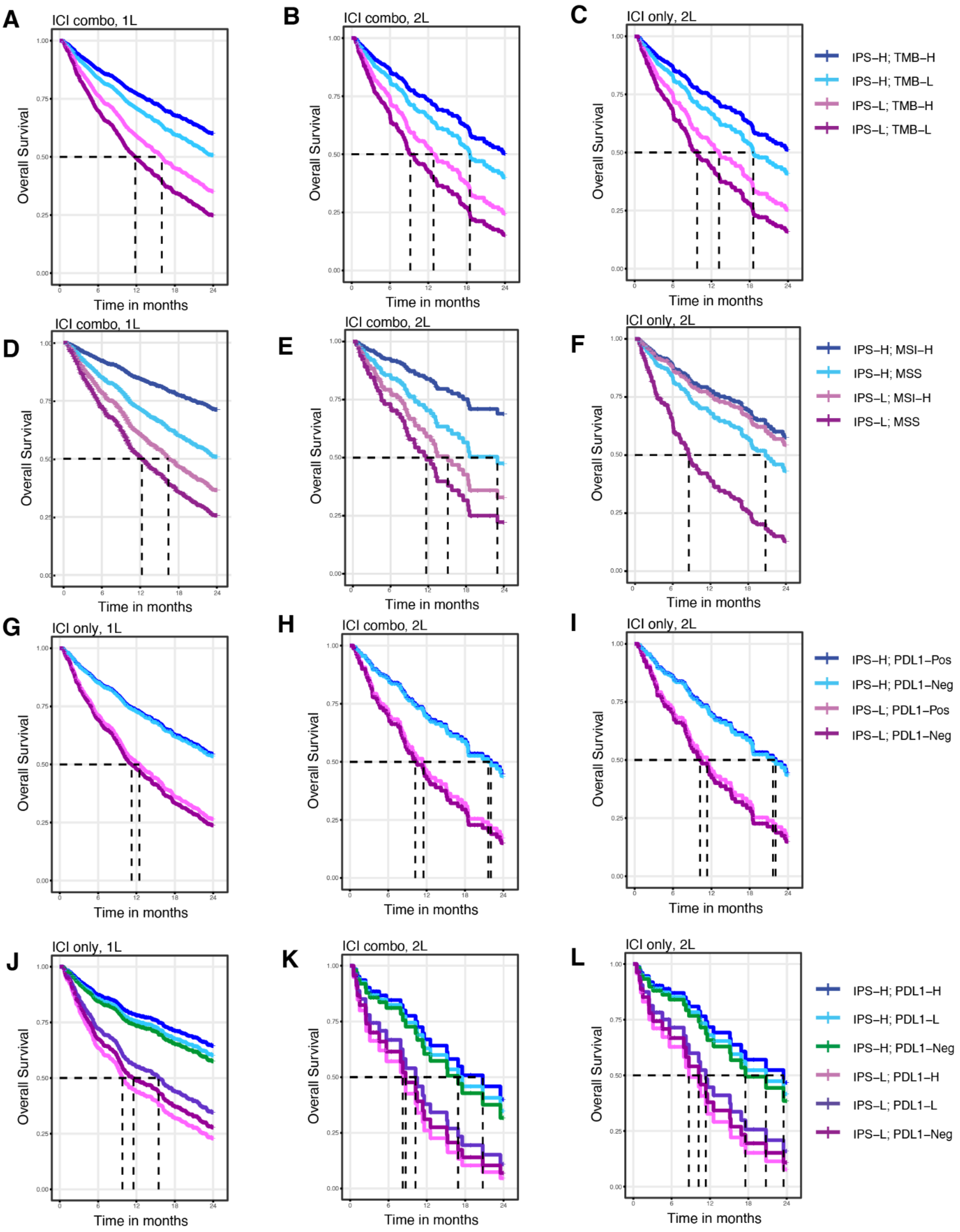
A
ICI combo, 1L
B
ICI combo, 2L
C
ICI only, 2L
D
ICI combo, 1L
E
ICI combo, 2L
F
ICI only, 2L
G
ICI only, 1L
H
ICI combo, 2L
I
ICI only, 2L
J
ICI only, 1L
K
ICI combo, 2L
L
ICI only, 2L
Overall Survival
Time in months
IPS–H; TMB–H
IPS–H; TMB–L
IPS–L; TMB–H
IPS–L; TMB–L
IPS–H; MSI–H
IPS–H; MSS
IPS–L; MSI–H
IPS–L; MSS
IPS–H; PDL1–Pos
IPS–H; PDL1–Neg
IPS–L; PDL1–Pos
IPS–L; PDL1–Neg
IPS–H; PDL1–H
IPS–H; PDL1–L
IPS–H; PDL1–Neg
IPS–L; PDL1–H
IPS–L; PDL1–L
IPS–L; PDL1–Neg

**Figure S2.**

Predicted OS curves from the CoxPH model described in **Figure 4b** for **a)** 1L combination therapy, **b)** 2L combination therapy, and **c)** 2L monotherapy patients. Predicted OS curves from the CoxPH model described in **Figure 4c** for **d)** 1L combination therapy,, **e)** 2L combination therapy, and **f)** 2L monotherapy patients. Predicted OS curves from the CoxPH model described in **Figure 4d** for **g)** 1L ICI-only therapy, **h)** 2L combination therapy, and **i)** 2L ICI-only therapy patients. Predicted OS curves from the CoxPH model described in **Figure 4e** for **j)** 1L ICI-only therapy, **k)** 2L combination therapy, and **l)** 2L ICI-only patients. All n values for these curves can be found in **Table S16**.

#### Figure S3

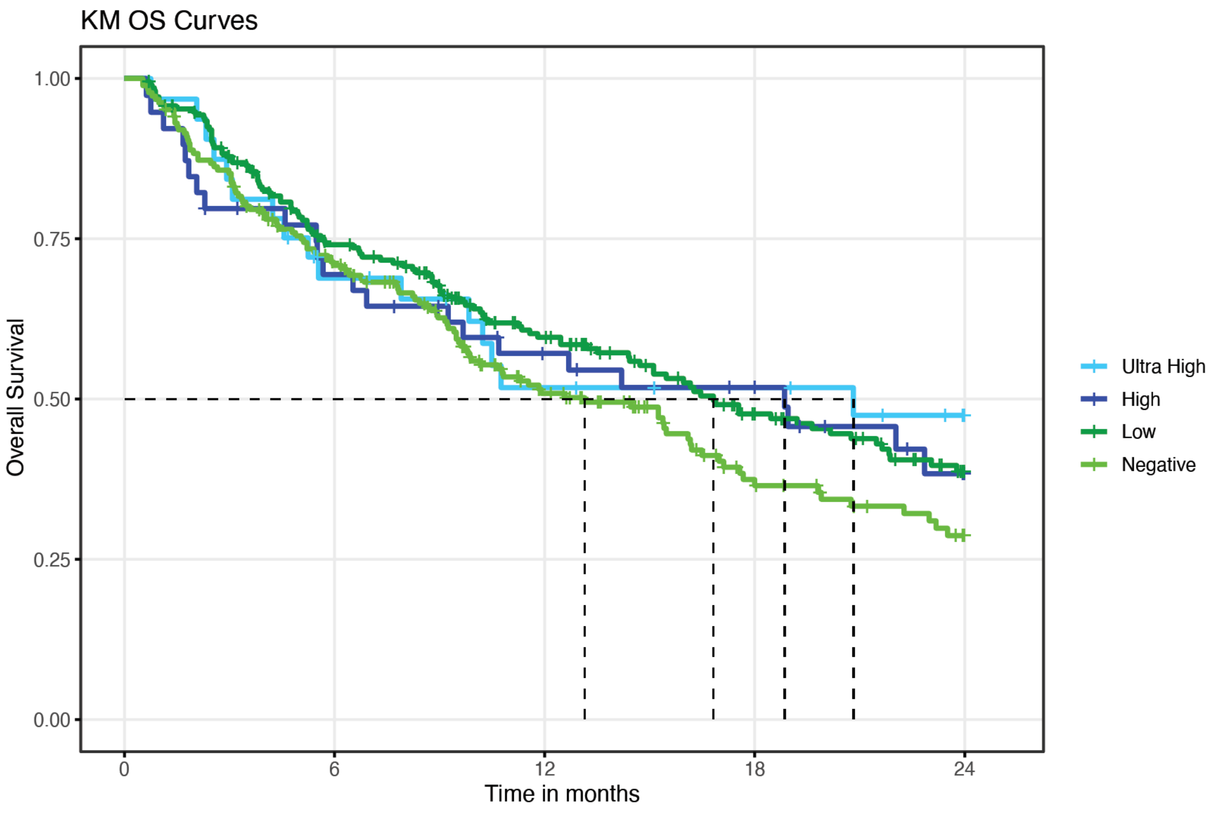


**Figure S3.**

KM Overall survival curves for NSCLC ICI+combination 1L cohort stratified by PDL1 IHC staining level. PD-L1 ultra high: TPS >90, PD-L1 high: TPS=89-50, PD-L1 low: TPS=49-1, PD-L1 negative: TPS=0.

**Figure S4**

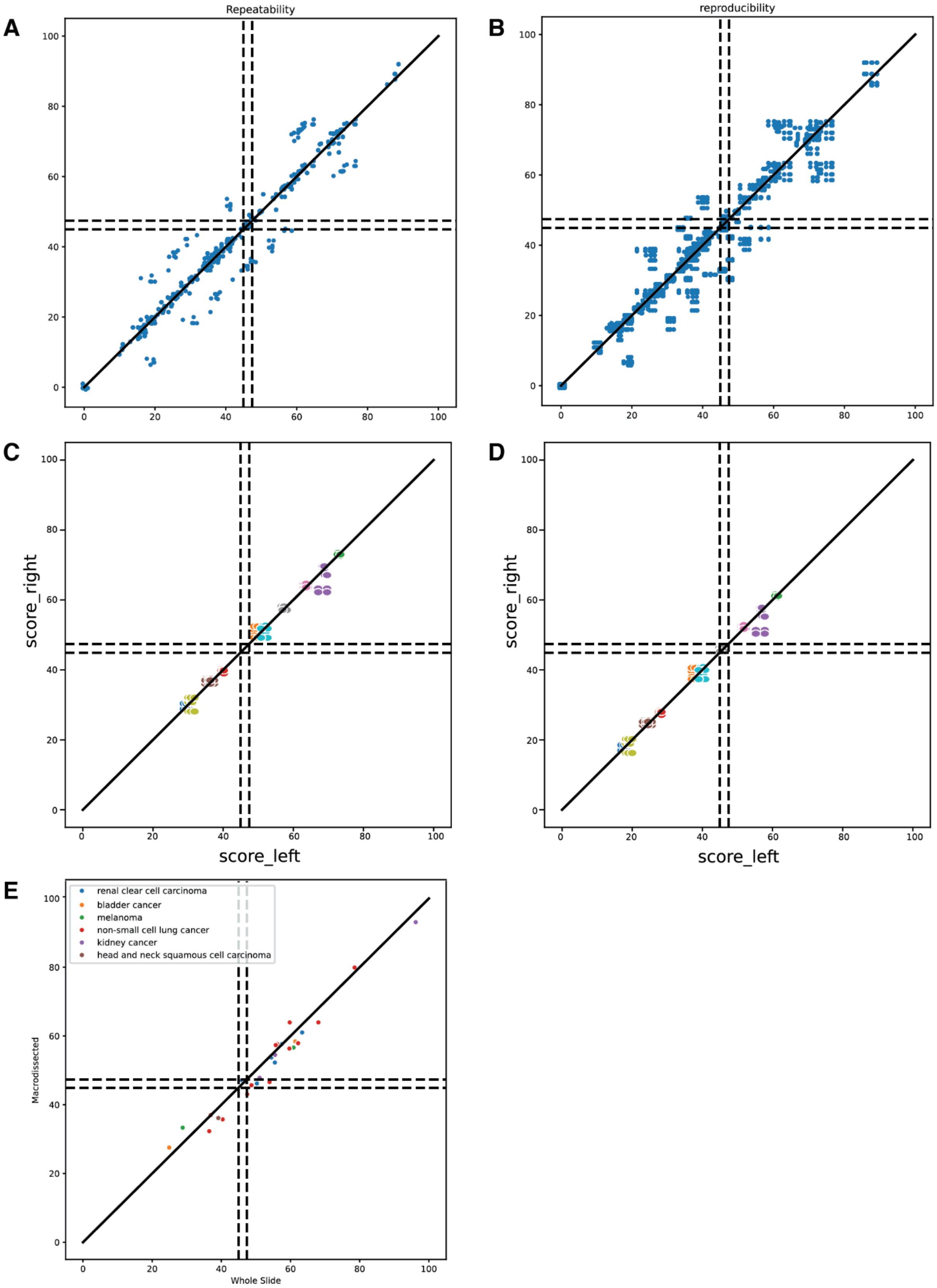
A
Repeatability
B
reproducibility
C
D
score_right
score_left
E
renal clear cell carcinoma
bladder cancer
melanoma
non-small cell lung cancer
kidney cancer
head and neck squamous cell carcinoma
Macrodissected
Whole Slide

**Figure S4.**

**a)** The repeatability of IPS was established by comparing the risk score and risk group of replicates performed under identical conditions.**b)** The reproducibility of IPS was established by comparing the risk score and risk group of replicates performed across different days and operators.The analytic sensitivity (mass input) was assessed by comparing the agreement of various mass input dilutions of a series of clinical RNA samples. Each clinical sample is grouped by color. **c)** Each RNA isolate is assigned a TMB-Low value and compared across each pair of replicates. **d)** Each RNA isolate is assigned a TMB-high value and compared across each pair of replicates. **e)** The analytic sensitivity of IPS is assessed using the macro-dissection status as a variable. Each sample is compared using both the macrodissected isolate and whole slide (non-macrodissected) isolate.
